# Pilot Clinical Trial to test the function of a Diagnostic Sensor in predicting Impending Urinary Catheter Blockage in Long-term Catheterized Patients

**DOI:** 10.1101/2022.10.25.22281351

**Authors:** Rachel A. Heylen, June Mercer-Chalmers, Annette Morton, James Urie, Edward Jefferies, Bethany L. Patenall, Maisem Laabei, A. Toby. A. Jenkins

**Affiliations:** Department of Chemistry, University of Bath, Claverton Down, Bath, BA2 7AY; Department of Urology, Royal United Hospital Bath NHS Foundation Trust, Combe Park, Bath, BA1 3NG; Mediplus Ltd, High Wycombe, UK; Department of Life Sciences, University of Bath, Claverton Down, Bath, BA2 7AY

## Abstract

**Trial design:** Pilot feasibility trial.

**Methods:** *Participants:* adults attending the Outpatient Urology Clinic, having a long-term indwelling urinary catheter and have the mental capacity to consent. Consent for the donation of the urinary catheter and drainage bag were gained at the Urology Clinic, Royal United Hospital (RUH) Bath, alongside a quality-of-life questionnaire.

*Interventions:* there was no direct intervention to the participants, the trial was to test the functionality of the diagnostic sensor, there was no change to the participant’s medical care.

*Objectives:* recruitment of 48 participants to donate catheter and drainage bags (including re-recruits); assess the functionality of the sensor to predict impending catheter blockage in human urine; assess the functionality to release at pH > 7; and assess the microbiological profile of the patients with long-term catheters.

*Outcome:* determination of whether the participant had a blockage event 3 weeks post catheter change, and whether this matched with the result from the sensor. Measurements of the participant’s urine to assess whether the sensor could detect human urine at a pH > 7. Determine the microbial species present in the drainage bag of the participants.

**Results:** *Recruitment:* received 35 samples from 28 individuals.

*Outcome:* two participants reported blockage events which were successfully detected by the sensor. However, the sensor also predicted blockage in participants that did not block within the 3-week report time period, sensitivity = 100%, specificity = 58.06%. The functionality of the sensor to detect urine above pH > 7 had a sensitivity = 78.75% and a specificity = 96.77%, which gave a *p* = 2.06×10^−24^ (χ^2^ test). Inclusion of the maintenance solution prescribed to participants, to test the predictability of the sensor, gave a sensitivity = 100%, and a specificity = 62.95%, *p* = 0.029 (Fisher Exact test). Microbiological analysis indicated that *Proteus* spp. and *Pseudomonas* spp. were the most commonly isolated microbes.

*Harm:* No adverse events.

**Conclusions:** The sensor can predict participants more prone to catheter blockage, and it is accurate in detecting urine with a pH >7. Owing to the small sample number of this trial, the results are not statistically powered. However, the data can be used to improve the design of the sensor and inform the design of a larger, randomized clinical trial.

**Trial registration:** Trial was ethically approved by the Research Ethics Committee (REC) number: 20/LO/0094. Integrated Research Application System (IRAS) number: 261095.

**Funding:** trial was funded by the Urology Foundation and an IAA seed grant, University of Bath.

## Introduction

A diagnostic sensor to detect impending catheter blockage has been developed as a medical device to aid long-term urinary catheter users. Urinary catheter blockage can occur as a results of urease-positive infections - the urease metabolizes urea to two molecules of ammonia and carbonic acid.^1^ This results in an increase in the pH within the bladder, causing struvite and apatite crystals to deposit onto and within the catheter, resulting in a crystalline biofilm that occludes the lumen of the catheter, leading to catheter blockage.^2,3^ Owing to the high rates of catheter-associated urinary tract infections (CAUTI), affecting approximately 150-250 million patients globally per year, urinary catheter blockage poses a significant problem in the clinic.^4^ The infections can remain within the bladder during catheter changes, form bladder stones, and migrate up towards the kidneys, causing kidney stones and pyelonephritis.^1,5^ In worst case scenarios, urosepsis can occur; ultimately patients experiencing urinary catheter blockage require more care, longer-stays in hospitals, and present a burden to the healthcare system.^6,7^

No device currently exists to predict catheter blockage. The sensor described in this study can be used by individuals with no existing medical knowledge. Placed within the catheter drainage bag, it provides a colorimetric indication of impending catheter blockage (Fig. 1), allowing the user or carer to change the catheter themselves; or flush the catheter using a maintenance solution thus reducing the risk of blockage.^8,9^ As this is a new medical device, a clinical trial is required to demonstrate efficacy. *In vitro* studies using a model of the catheterized tract have shown that the sensor provides 6.7 h warning prior to catheter blockage.^9^ Having shown to be stable within healthy human urine, the next stage was to test the sensor in urine taken from long-term catheterized patients.^9^ A small-scale pilot study was set up, in collaboration with the Department of Urology, RUH; participants donated their drainage bag and catheter, allowing assessment of the sensor. Here we report, the results of the feasibility study to test a diagnostic sensor to detect impending urinary catheter blockage.

**Figure 1.**
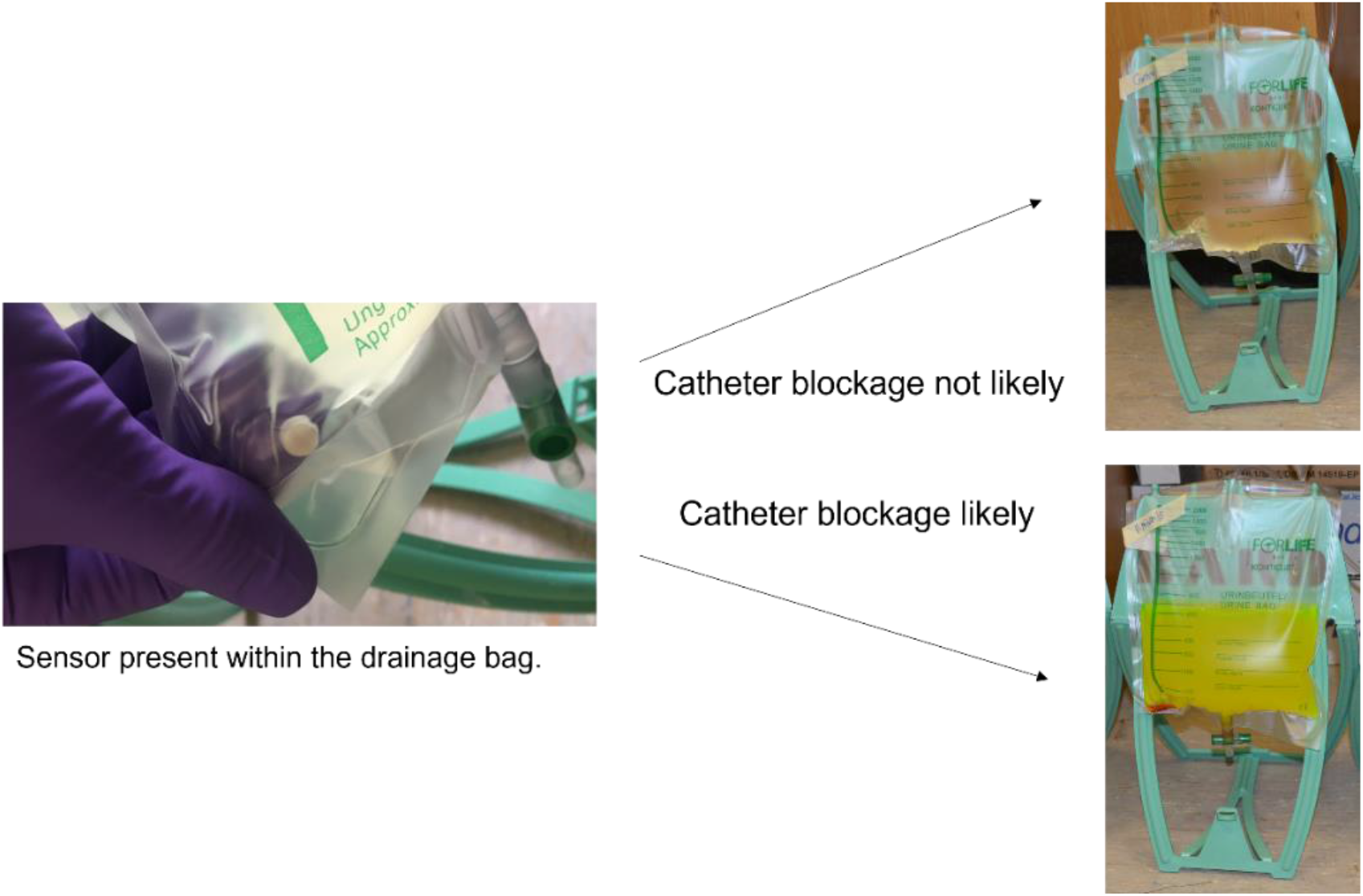
Diagram showing the colorimetric change when the sensor indicates that catheter blockage is likely to occur.^9^

## Participants and Methods

### Participants

Eligible participants had to be ≥18 yr. and have the mental capacity to consent to the study. They also had to use an indwelling long-term urinary catheter which would be changed at the Outpatient Urology Clinic, RUH. During recruitment, the participants were asked to complete a quality-of-life (QoL) survey - an International Consultation on Incontinence Questionnaire (ICIQ) developed by Cotterill *et al*., specifically for long-term urinary catheter users (Supplementary Document 1).^10^

### Study Design

The study had two aims: (1) to determine the feasibility for a larger multi-center trial to test the diagnostic sensor, (2a) assess the predictability of the sensor - does sensor turn on correlate with reported catheter blockage; (b) assess the functionality of the sensor - does it release at pH > 7; and (c) investigate the microbial diversity of the drainage bag.

The study was voluntary, participants were offered a written leaflet describing the study (Supplementary Document 2) and allowed to ask any questions of the research clinical staff, prior to written consent being gained. A screening process took place to determine whether the donation could be taken, and a clinical record form was completed (CRF) (Supplementary Document 3). Participants donated their full (>150 mL) urine drainage bag and their catheter. The samples were blinded and stored between 2-8 °C before pick-up and transport to University of Bath for analysis. There was no change to the treatment plan of the participants. Three weeks after donation, a follow up telephone call was completed to determine if a blockage event had occurred or if there had been a change in the catheter care plan (Supplementary Document 4). Owing to time, staff restraints and the capacity at the RUH, it was determined that 48 participant samples would be a suitable number for a small-scale feasibility trial.

### Sample Processing

All materials were purchased from ThermoScientific, UK, unless otherwise stated. The samples were blinded at the RUH: once the samples were collected, there was no identifiable link from the sample to the participant at the University of Bath. CRF containing the medical information of the participants was analyzed after the samples had been processed; to prevent biases between the laboratory results and the contents of the CRF. Samples were stored between 2-8 °C until processing. All analysis of the urine was conducted using aseptic techniques within a sterile microbiological cabinet. Each sample contained the drainage bag and the catheter, both were photographed allowing a record of the color of the urine and whether the tip of the catheter was encrusted. Urine from the drainage bag was aliquoted into six separate 50 mL falcon tubes (Corning), 10 mL per tube. Positive and negative controls were prepared using artificial urine, prepared according to Nzakizwanayo *et al*., for the positive control 1 M NaOH was added to buffer the artificial urine to a pH > 8.^11^ A sensor was dropped into five of the six tubes containing the participant’s urine, one tube was left as a no sensor control. The tubes for one participant, including the positive and negative control (also containing sensors) were photographed in a light box, sample distance 30 cm, and with the same camera setting each time (Nikon D3100, 35 mm lens zoom) (Supplementary Figure 1). For three of the tubes the pH and temperature were measured, then averaged. The tubes for all participants were incubated at room temperature for 18 h. After the incubation, the tubes were photographed as previously described, the release of the sensor was visually determined. The tubes were re-measured for pH and temperature, as described.

### Microbial analysis

A 10 µL loop of urine was streaked, to attain individual colonies to assess morphology, onto the following agar plates: Cystine-Lactose-Electrolyte Deficient (CLED), MaConkey (MC), Müeller Hinton (MH), Lurie-Bertani (LB), non-swarming LB (NSLB, tryptone (10 g/L), yeast extract (5 g/L) and bacteriological agar (15 g/L)), Columbia blood agar (CBA) (5% sheep blood) and CHROMID agar plates (bioMérieux, UK). An additional CBA plate was prepared by streaking 10 µL loop using the semi-quantitative streaking method. All plates were statically incubated overnight at 37 °C. Following incubation, the plates were photographed using a light box, and the bacteria were visually described by determining the color, size of colonies, and whether the colonies had a smooth or rough edge. The semi-quantitative plate was compared to a key containing known concentrations of bacterial growth (Supplementary Document 6). For polymicrobial samples, the individual colonies were picked and re-streaked to attain separate, single colonies. From the plates, individual colonies were picked and grown in either LB media or MH media from which freezer stocks were prepared containing 15 % (v/v) glycerol, and placed in at -80 °C for long-term storage. Each of the distinctive colonies was tested for urease activity using the phenol red method, a small tip of a colony was mixed with phenol red solution (10% (w/v) urea and 0.02% (w/v) phenol red), positive result was reported if the color changed from yellow to red. A tentative assessment on the identification of the bacterial species was made from the urease activity assay and the appearance of the bacterial colonies on the selective agar plates (MC, CLED, and CHROMID). Figure 2 shows a schematic of the study design.

**Figure 2.**
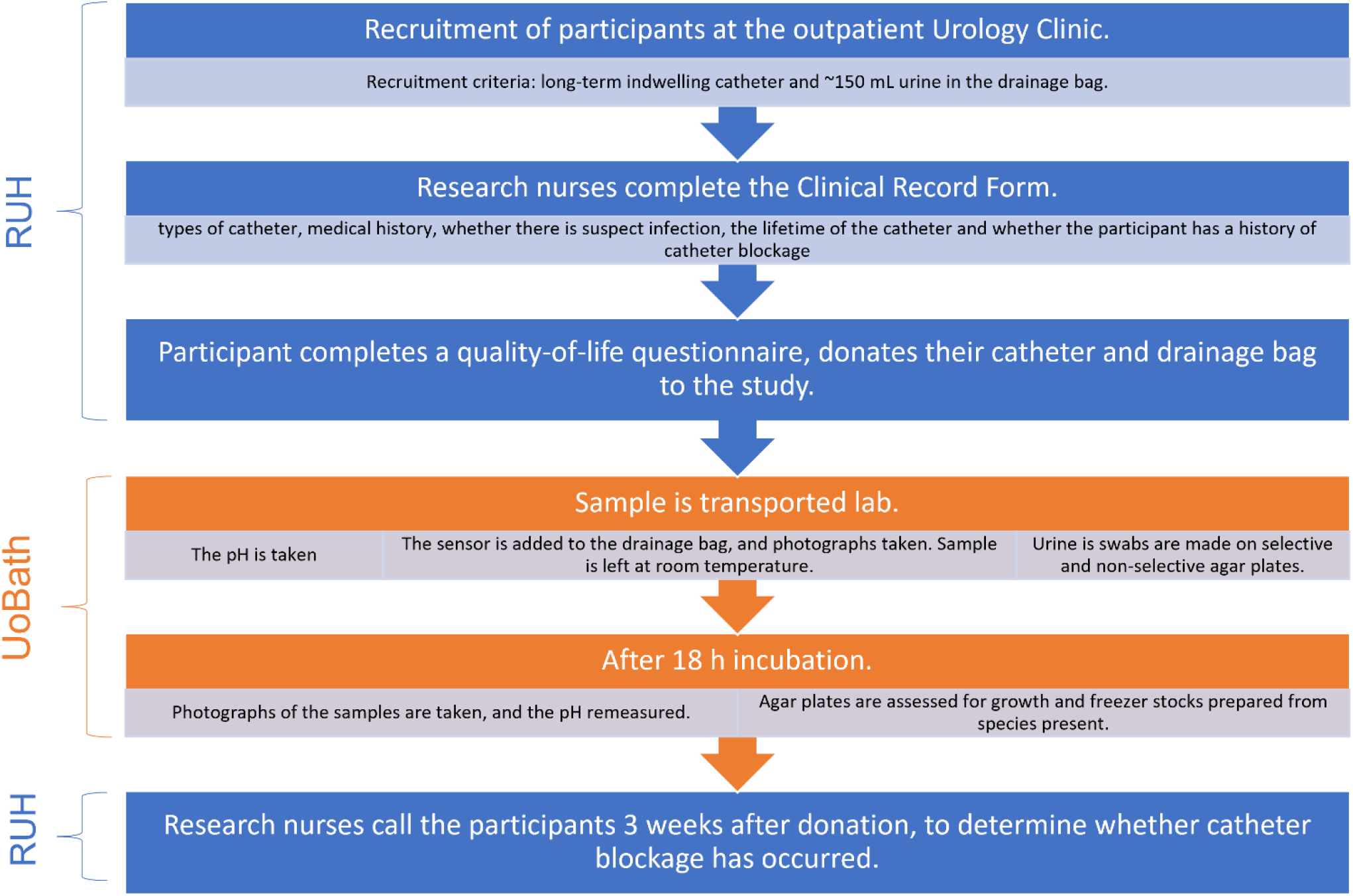
Schematic showing the study design and the processing of the samples from the Royal United Hospital (RUH) to the University of Bath (UoBath).

### 16S rRNA sequencing

Identification of the bacteria was confirmed using 16S rRNA sequencing. To achieve single colonies, the bacteria were grown, statically at 37 °C overnight, from the freezer stocks onto LB plates. Preparation for colony PCR was conducted within a sterile cabinet, to prevent amplification of other bacteria as these primers are universal (Table 1). Primers were diluted to 100 µM using nuclease-free water (Sigma). From the plates, a single colony was picked, added to 100 µL of nuclease-free water, and microwaved on high for 3 min. Within an autoclaved PCR tube (Greiner), 2 µL of colony preparation was added, along with 18 µL of master mix. Master mix contained (per reaction) 10 µL PHUSION high-fidelity enzyme mix, 1 µL 27F primer, 1 µL of 1392R primer, and 6 µL of nuclease-free water. PCR was run using a PHUSION method; melting temperature of 56 °C, and extension time of 1 min for 35x cycles at 72 °C. The resulting product was run on a 1% (w/v) agarose gel at 110 V for 30 min, a 1 kb ladder was used to confirm the PCR product. If the colony PCR failed for any of the strains, then the DNA was extracted using a Sigma DNA extraction kit. As these were unknown strains, if the Gram positive/negative could not be determined from the tentative assessment made from the selective plate analysis, then the DNA was extracted using both the Gram-positive and Gram-negative methods. The PCR product was cleaned using a Wizard SV Gel and PCR Clean-up System (Promega). The concentration of the DNA was determined using spectrometer, A_260_, and diluted to a final concentration of 10 ng/µL. Two samples were prepared per strain, 2 µL of the 27F primer was added to one sample and the 1392R to the other. This allowed sequencing in both directions across the PCR product, the maximum read length for the sequencing was 1000 bp, however the 16S rRNA sequence amplified was ∼1500 bp. The prepared solutions were sequenced by Eurofins, the quality of the sequence was analyzed, and the sequences were trimmed to remove less reliable base reads. The sequences were run through the EzBioCloud Database and the identification analyzed. Both reads were run separately, and the results compared to the initial tentative assessment.

**Table 1.**
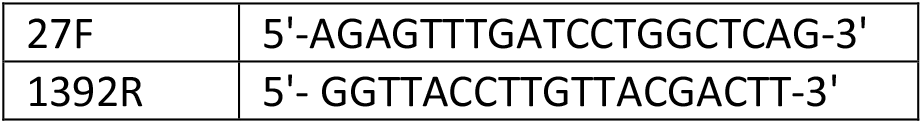
Primers used for the PCR, to amply the 16S rRNA sequence. ^12^

### Statistical Analysis

This was a small-scale pilot study, therefore there was no randomization, and the study was not powered. The following statistical tests were used to determine significance: Fisher Exact test and the χ^2^ test.

## Results and Discussion

### Recruitment

The study recruited 35 samples, from 28 individuals. The study aimed to obtain 48 samples, however, owing to staffing restrictions at the RUH, it was decided to stop collection after the 35 samples. The trial ran over 5 months and recruitment occurred at 15 outpatient clinics (average of 2.3 participants recruited/clinic). The main reason participants were not recruited was because the drainage bags did not have > 150 mL of urine. No samples were excluded from the study. Table 2 indicates the baseline demographics and clinical characteristics. Supplementary Table 1 summaries the different catheter manufacturers. In the study, the majority of participants were male (82.1%). Shackley *et al*., reported a 3:2 ratio in male vs females who were catheterized (total number catheterized 1 194 902), this ratio is not reflected in this study.^13^ Shackley *et al*. included patients with short-term catheters as well as long-term, and therefore the 3:2 ratio might not be representative of long-term users. The average age of all participants was 72.6 yr., all participants were white in ethnicity, the demographic in Bath and North-East Somerset Trust, UK, is 90% White: British, therefore for a small sample size this result is expected.^14^

**Table 2.**
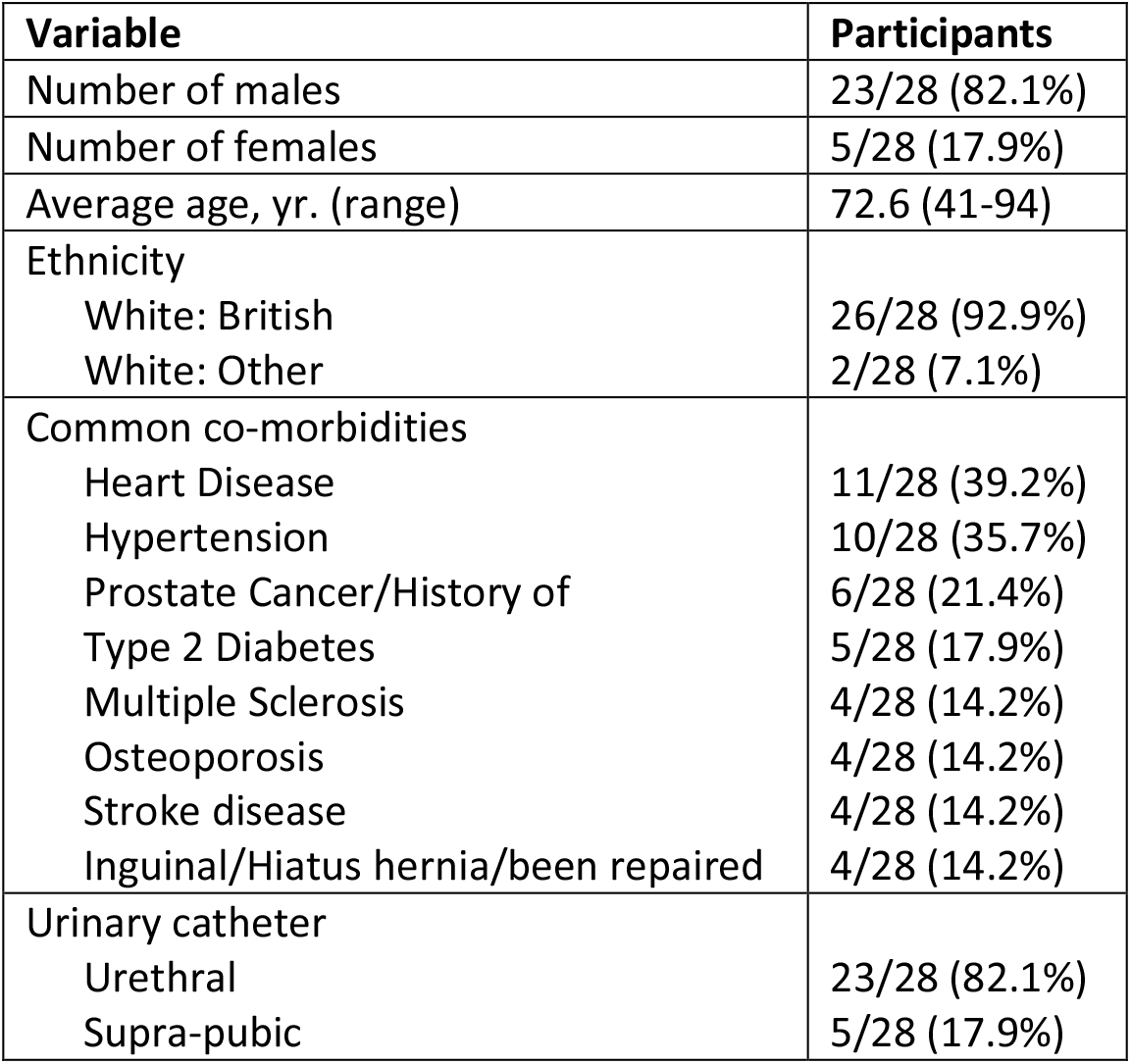
The baseline demographics and clinical characteristics of the participants in the study.

| Variable | Participants |
| --- | --- |
| Number of males | 23/28 (82.1%) |
| Number of females | 5/28 (17.9%) |
| Average age, yr. (range) | 72.6 (41-94) |
| Ethnicity |  |
| White: British | 26/28 (92.9%) |
| White: Other | 2/28 (7.1%) |
| Common co-morbidities |  |
| Heart Disease | 11/28 (39.2%) |
| Hypertension | 10/28 (35.7%) |
| Prostate Cancer/History of | 6/28 (21.4%) |
| Type 2 Diabetes | 5/28 (17.9%) |
| Multiple Sclerosis | 4/28 (14.2%) |
| Osteoporosis | 4/28 (14.2%) |
| Stroke disease | 4/28 (14.2%) |
| Inguinal/Hiatus hernia/been repaired | 4/28 (14.2%) |
| Urinary catheter |  |
| Urethral | 23/28 (82.1%) |
| Supra-pubic | 5/28 (17.9%) |

### QoL responses

The QoL questionnaire was completed by 28/28 (100%) participants in the study. The catheter function and concerns score, 11.5, was below the reported mean score by Cotterill *et al*., of 14.5 (n = 199) however it was within ± 1 standard deviation (Table 3).^10^ Similarly, the lifestyle impact score, 7.0, was below the reported mean score of 8.6 (n = 202) but within ± 1 standard deviation. Therefore, the QoL questionnaire gave results comparable to previously reported data.^10^ Owing to the focus of this study, Q6, which related to urinary catheter blockage, was particularly important. Generally, there was a low score for this question, suggesting that the participants were not concerned about catheter blockage (Table 3).

**Table 3.** Scores and ranges for the quality-of-life questionnaire, calculated as described by Cotterill *et al*.^10^ Table 3 additionally shows the scores for Q6 which relates specifically to urinary catheter blockage.

| Domain | Score |
| --- | --- |
| Catheter function and concerns (n = 28) | Range = 1-23, mean = 11.5 |
| Lifestyle impact (n = 28) | Range = 3-15, mean = 7.0 |
| Q6a: 'Is the possibility of the catheter blocking on your mind?' (n = 27) | Range = 0-4, mean = 1.04 |
| Q6b: 'How much does this bother you?' (n = 27) | Range = 0-10, mean = 2.44 |

### Sensor performance

The sensor performance was measured by the turn-on of fluorescence observed in the 18 h incubation period and whether this correlated with a reported blockage event (Fig. 3A). From the study, only two patients had a blocked catheter within the three-week follow-up phone call. As these catheters are designed to be *in situ* for up to three months, it is likely that, even for recurring “blockers”, no blockage may have been expected within the three-week lag time between collection and follow-up phone calls. The sensitivity was recorded at 100%, the sensor correctly predicted the participants which did block. However, the specificity was 58.06%, this is due to the large number of sensor turn-on events when the catheters did not block (false positive results).

**Figure 3.**
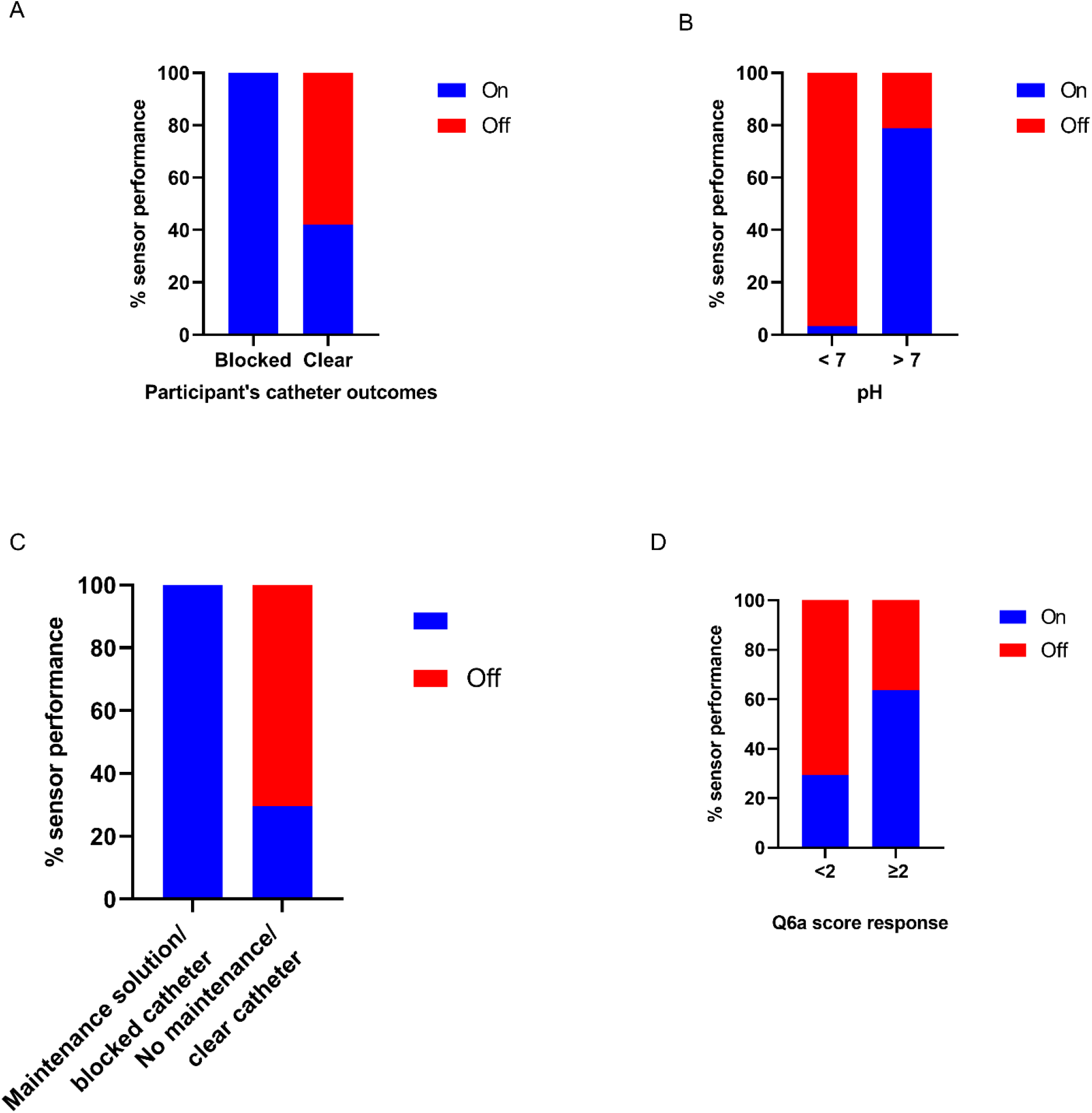
**A**. Predictability of the sensor determined by the number of sensors which turned on compared to the reported blockage events. **B**. The functionality of the sensor determined by the number of sensors turned on compared to the measured pH. **C**. The predictability of the sensor determined by number of sensors which turned on compared to the reported blockage events and prescribed maintenance solutions. **D**. Comparison of the sensor performance with the concern for catheter blockage, as reported in Q6a of the quality-of-life questionnaire. Graphs were prepared using GraphPad Prism v. 9.4.1.

The functionality of the sensor was tested by measuring the pH of the urine and correlating it with sensor turn-on in the 18 h incubation (Fig. 3B). It is well documented in the literature that urease activity increases the pH of the urine, leading to an increased likelihood of catheter blockage.^3,15,16^ Long-term catheter users who have reoccurring blockages (blockers) are more likely to have a void urine pH > 7.^15,17,18^ The 173 sensors tested reported a sensitivity of 78.75% and a specificity of 96.77%. Therefore, the sensors demonstrated strong evidence that they turn-on at pH > 7 (*p* = 2.06 × 10^−24^, χ^2^ test).

Within the CRF (Supplementary Document 3), Q31-32, records the use of flush out solutions or maintenance solutions. Flush out solutions generally are saline or water, they are flushed up the catheter and are designed to remove debris. Maintenance solutions are citrate based and are designed to buffer the pH within the bladder. Current evidence that these solutions are effective is poor, however they are still prescribed to frequent blockers.^19^ From the samples received, three individuals (six samples) were prescribed washout/maintenance solutions. As these are only used for patients who are frequent blockers, these were added to the predictability test result (Fig. 3C). With the addition of these results, the sensitivity remained at 100% and the specificity improved to 70.37%. Using a Fisher Exact test, there is significance between being prescribed a maintenance solution/reporting a blocked catheter and sensor turn-on (*p* = 0.029). There was no correlation in the time between catheter changes and sensor turn-on, however, all participants who reported blockages, or were prescribed maintenance solutions had a pH > 7 (except RUH07 = 6.73, raising to 7.44 after 18 h incubation) (average is 8.20, range 6.73-8.80).

Considering the participant’s concerns about urinary catheter blockage (Q6, Supplementary Document 1), a correlation is observed between sensor turn on and those participants who expressed a concern about blockage (<2 (never (0), occasionally (1)) and ≥2 (somewhat (2), most-of-the-time (3), all-of-the-time (4)). Table 3 showed that the mean of those concerned with catheter blockage was at 1.04. However, if a threshold of ≥2 is taken, sensor turn-on is associated with these participants. Figure 3D, shows the sensor performance relating to the score to Q6a. A sensitivity of 70.59% and a specificity of 63.64% was calculated here for the performance of the sensor relating to the participants concerns about catheter blockage. This result suggests that participants who were concerned about catheter blockage were more likely to get sensor turn-on, although this does not link directly to the physiological blockage events, nor the high pH of the urine. If the participants recorded a concern for blockage, it is likely they have experienced the symptoms of blockage before, therefore potentially making it more likely for them to be blockers. It is reported in the literature that once a patient experiences blockage, they are more likely to become a recurrent blocker, owing to the likelihood of the urease-positive infection remaining within the bladder between catheter changes.^15,16^

### Microbial analysis

A total of 90 strains were isolated from participant’s urine. These underwent tentative assignment using selective plates. Owing to redundancy identified by the streaked plates analysis, 71 were sent for 16S rRNA analysis. The following strains failed during the sequencing step: RUH07-01, RUH14-02, RUH16-01, RUH20-02, and RUH30-01. Each of these was attempted three times, therefore their identification was made from the selective plate analysis, except for RUH30-01 which was grown on a non-selective CBA plate and was not identified. From the 35 samples analyzed, 22/35 (62.9%) were polymicrobial and 13/35 (37.1%) had one species present. No urine samples were completely bacteria-free, despite 33/35 participants reporting that they did not suspect an infection, suggesting the presence of an asymptomatic infection (Q2, CRF, Supplementary Document 3). This agrees with reports that long-term catheterized patients have chronic bacteriuria, often asymptomaticly.^16^ Contamination by skin commensals during the removal of the catheter could have occurred however, this is unlikely as the nurses are using aseptic technique for catheter removal and replacement. Table 4 indicates the frequency of the species identified from the 35 samples, full details of the bacteria identified for each individual sample can be found in Supplementary Table 2.

**Table 4.** Frequency of the bacterial species from the samples. ^*^Four of the *Escherichia spp*. were identified as *Shigella flexneri*, these have been reported as being closely related. Although distinct species here, they have been grouped together to allow comparison to other analysis of long-term CAUTI.^20^

| Bacteria species | Frequency | Urease activity | Reference |
| --- | --- | --- | --- |
| *Proteus spp.* | 13 | Positive | 3 |
| *Pseudomonas spp.* | 10 | Variable | 21 |
| *Enterococcus spp.* | 10 | Negative | 22 |
| *Klebsiella spp.* | 9 | Positive | 23 |
| *Escherichia spp.* * | 8 | Negative | 24 |
| *Staphylococcus spp.* | 4 | Variable | 24 |
| *Citrobacter koseri* | 3 | Variable | 25 |
| *Enterobacter spp.* | 2 | Variable | 26 |
| Other | 5 | N/A |  |

Examination of the relationship between the urease activity and those that reported catheter blockage, or the use of maintenance solutions, showed that all those with an increased likelihood of blockage had a urease positive infection, in agreement with literature (Fig. 4A).^1,3,5–7,27^ However, urease activity was measured in just over half the samples, with no indication of an increased blockage likelihood. Within patients, there are complex interactions between the bacterial species present and between bacteria and the patient. It can be hypothesized that, owing to many of these samples containing polymicrobial infections, there could be intra-species competition within the bladder which is repressing the urease positive bacteria. Or conversely, as reported by Armbruster *et al*., urease activity could be enhanced by the presence of different species, such as the synergy observed between *P. mirabilis* and *Providencia stuartii* which leads to enhanced urease activity.^2,27^ Additionally, the participant’s immune system may also limit infection severity by controlling bacterial numbers below an infection threshold. As this study has a low sample size, larger studies are required to fully investigate these interactions. One of the limitations of this experiment is the bacteria have not been quantified, with no data on the quantities of the bacteria within the urine. This was not completed as freezing urine, which would have allowed for quantitative analysis, was not possible at the time, also to fully understand the quantity of bacteria within the bladder sterile samples of urine from the bladder should be taken, not from the drainage bag as this reflects a different environment. Another limitation is that only aerobic bacteria were detected, owing to the limitations in the methodology. Anaerobic bacteria could have been present and competing with the bacteria present. Despite this limitation, the identification of the bacteria here is more rigorous than standard clinical testing. The sensor performed well in detecting the urease positive infections, with a measured sensitivity of 63.64% and a specificity of 84.62% (Fig. 4B). Interestingly, in two of our blocked/maintenance participants had both *P. mirabilis* and *Enterococcus faecalis* infections, Gaston *et al*., reported that there was polymicrobial interactions between these species which facilitated biofilm formation, antibiotic recalcitrance and increased the persistence within the catharized urinary tract.^28^

**Figure 4.**
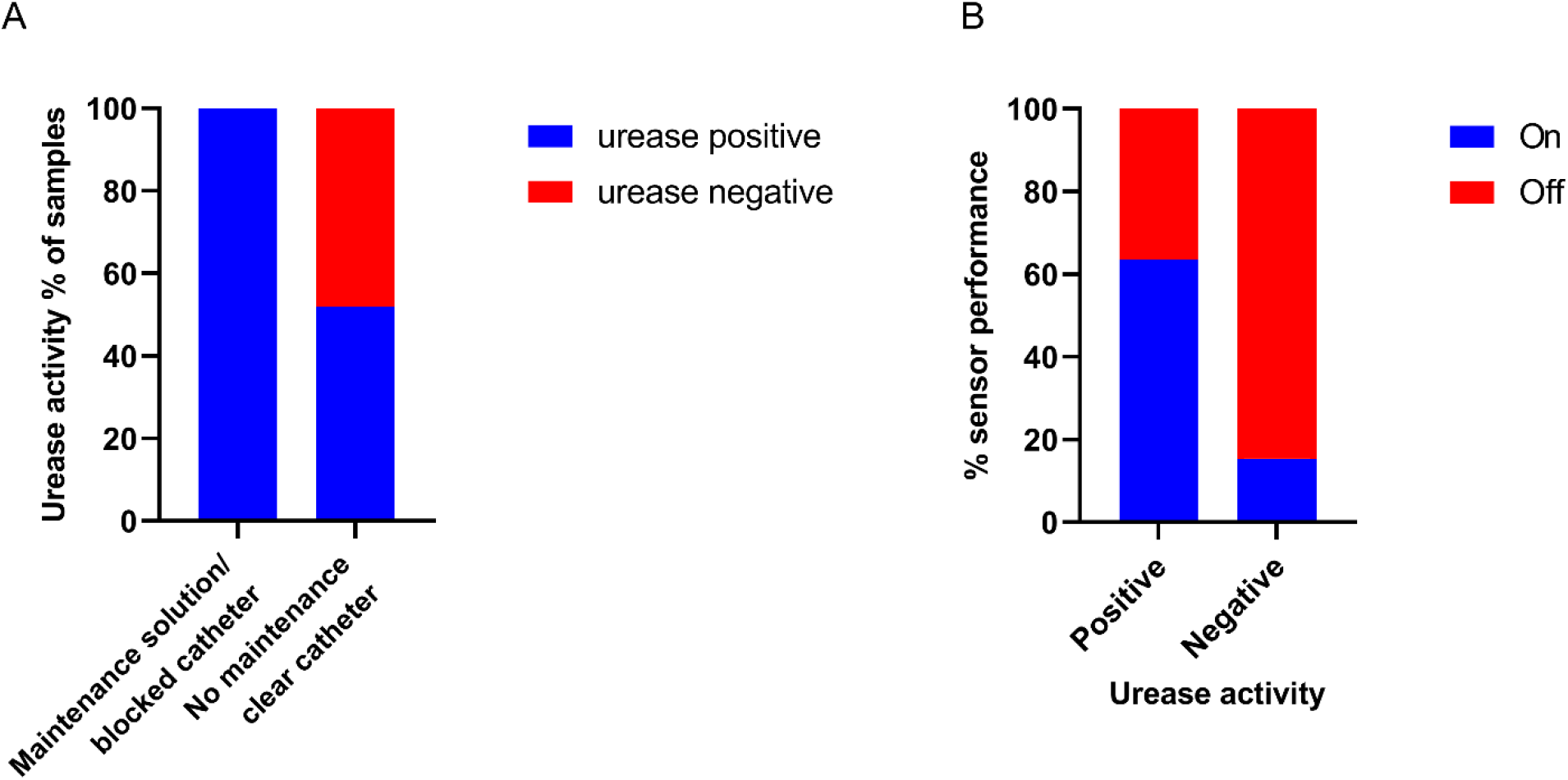
**A**. Relationship between urease activity and reported blockages or the use of maintenance solution. **B**. Correlation between sensor turn on and measured urease activity. Graphs were prepared using GraphPad Prism v. 9.4.1.

## Discussion

### Limitations

The major limitation of the study is the sample size, which is small and lacks variation in the baseline characteristics (Table 1). As a result of the sample size, limited statistical analysis could be carried out on the ability of the sensor to detect impending blockage. The second limitation of the study is the delay time between the change of catheter and the follow-up telephone call (3 weeks). This discrete timepoint of 3 weeks prevents further follow up of the participants and restricts the results of the study. Only two participants reported a blockage within that timeframe. In the planning of a larger clinical trial, these limitations can be avoided, a larger sample size will be used; the sensor shall be placed within the drainage bag and given to the participants for use over a continuous time-period. Participants could then frequently report their sensor status (remotely using an app) allowing constant monitoring of the sensor.

### Sensor performance

The sensor accurately predicted the two blockage events which occurred, it also was able to detect participants who have an increased likelihood of catheter blockage (prescribed maintenance solutions) and those who’s samples tested urease positive in the urease activity assay (Fig. 3A&C, 4B). However, it did report several false-positive results where the sensor turned on, but the catheter did not block. It is likely that future use of the sensor will require a two-fold threshold to detect impending blockage: (1) sensor turns on in drainage bag, (2) how the patient is feeling, increased frequency in the use of maintenance solutions/reduced urine output. This will require future testing in a larger clinical trial.

### Microbiological analysis

Despite urinary catheters being frequently supplied to patients, there are limited studies investigating the bacteria within long-term urinary catheter users. Indeed, most studies that have incorporated microbial analysis have a small sample size, similar to this study.^15,29–31^ Species identified in this study are similar to those observed in previous studies - these studies also have the similar limitation of only identifying aerobic bacteria. Two of the studies used a MacConkey and blood agar streak method.^15,30^ Whilst the others used standard clinical microbiology, which in the UK consists of CLED streaking. Our methodology incorporated both selective agar plating and 16S rRNA sequencing. This more investigative approach identified the species-level differences between *E. coli* and *S. flexneri*, which are not observed with selective plate methods. Indeed, the selective agar plating mis-identified 50 strains sent for sequencing, indicating the importance of sequence level identification.

### Conclusions

This pilot small-scale clinical study, conducted with long-term urinary catheter participants, demonstrates the use of a sensor to diagnose impending urinary catheter blockage. The sensor accurately identified the blockage events occurring in the study. However, owing to low sample numbers, statistical analysis could not be completed. This pilot study will be used to design a larger scale study to test the sensor.

## Data Availability

All data produced in the present study are available upon reasonable request to the authors

## References

1. Norsworthy AN, Pearson MM. From Catheter to Kidney Stone: The Uropathogenic Lifestyle of Proteus mirabilis. Trends Microbiol. 2017;25(4):304–315. doi:10.1016/j.tim.2016.11.015

2. Armbruster CE, Mobley HLT, Pearson MM. Pathogenesis of Proteus mirabilis Infection. EcoSal Plus. 2018;8(1):1–73. doi:10.1128/ecosalplus.esp-0009-2017

3. Stickler DJ, Feneley RCL. The encrustation and blockage of long-term indwelling bladder catheters: A way forward in prevention and control. Spinal Cord. 2010;48(11):784–790. doi:10.1038/sc.2010.32

4. Zowawi HM, Harris PNA, Roberts MJ, et al. The emerging threat of multidrug-resistant Gram-negative bacteria in urology. Nat Rev Urol. 2015;12(10):570–584. doi:10.1038/nrurol.2015.199

5. Choong S, Wood S, Fry C, Whitfield H. Catheter associated urinary tract infection and encrustation. Int J Antimicrob Agents. 2001;17:305–310. doi:https://doi.org/10.1016/s0924-8579(00)00348-4

6. Schnauffer JN, Pearson MM. Proteus mirabilis and Urinary Tract Infections. Microbiol Spectr. 2015;3(5):1032–1057. doi:10.1111/mec.13536.Application

7. Getliffe K, Newton T. Catheter-associated urinary tract infection in primary and community health care. Age Ageing. 2006;35(5):477–481. doi:10.1093/ageing/afl052

8. Milo S, Acosta FB, Hathaway HJ, Wallace LA, Thet NT, Jenkins ATA. Development of an Infection-Responsive Fluorescent Sensor for the Early Detection of Urinary Catheter Blockage. ACS Sensors. 2018;3(3):612–617. doi:10.1021/acssensors.7b00861

9. Heylen RA, Branson M, Gwynne L, et al. Optimization of a Lozenge-Based Sensor for Detecting Impending Blockage of Urinary Catheters. Biosens Bioelectron.:1–11.

10. Cotterill N, Fowler S, Avery M, et al. Development and Psychometric Evaluation of the ICIQ-LTCqol: A Self-Report Quality of Life Questionnaire for Long-Term Indwelling Catheter Users. Neurourol Urodyn. 2016;35:423–428. doi:10.1002/nau.22729

11. Nzakizwanayo J, Pelling H, Milo S, Jones B V. An In Vitro Bladder Model for Studying Catheter-Assoicated Urinary Tract Infection and Asssoicated Analysis of Biofilms. (Pearson M, ed.).; 2019.

12. Srinivasan R, Karaoz U, Volegova M, et al. Use of 16S rRNA gene for identification of a broad range of clinically relevant bacterial pathogens. PLoS One. 2015;10(2):1–22. doi:10.1371/journal.pone.0117617

13. Shackley DC, Whytock C, Parry G, et al. Variation in the prevalence of urinary catheters: A profile of National Health Service patients in England. BMJ Open. 2017;7(6):1–8. doi:10.1136/bmjopen-2016-013842

14. Cox T. Bath and North East Somerset Clincial Commissioning Group. https://bsw.icb.nhs.uk/wp-content/uploads/sites/6/2022/06/Annual-Report-BaNES-CCG-2018-19-FINAL-1.pdf.

15. Kunin CNM. Blockage of urinary catheters: Role of microorganisms and constituents of the urine on formation of encrustations. J Clin Epidemiol. 1989;42(9):835–842. doi:10.1016/0895-4356(89)90096-6

16. Nicolle LE. Catheter associated urinary tract infections. Antimicrob Resist Infect Control. 2014;3(1):1–8. doi:10.1186/2047-2994-3-23

17. Choong SKS, Hallson P, Whitfield HN, Fry CH. The physicochemical basis of urinary catheter encrustation. Br J Urol Int Int. 1999;83(7):770–775. doi:10.1046/j.1464-410x.1999.00014.x

18. Mathur S, Suller MTE, Stickler DJ, Feneley RCL. Prospective study of individuals with long-term urinary catheters colonized with Proteus species. Br J Urol Int. 2006;97(1):121–128. doi:10.1111/j.1464-410X.2006.05868.x

19. Hagen S, Sinclair L, Cross S. Washout policies in long-term indwelling urinary catheterisation in adults. Cochrane Database Syst Rev. 2017;2017(3). doi:10.1002/14651858.CD004012.pub5

20. Devanga Ragupathi NK, Muthuirulandi Sethuvel DP, Inbanathan FY, Veeraraghavan B. Accurate differentiation of Escherichia coli and Shigella serogroups: challenges and strategies. New Microbes New Infect. 2018;21:58–62. doi:10.1016/j.nmni.2017.09.003

21. Bradbury RS, Reid DW, Champion AC. Urease production as a marker of virulence in Pseudomonas aeruginosa. Br J Biomed Sci. 2014;71(4):175–177. doi:10.1080/09674845.2014.11978060

22. García-Solache M, Rice LB. The enterococcus: A model of adaptability to its environment. Clin Microbiol Rev. 2019;32(2). doi:10.1128/CMR.00058-18

23. Mobley HLT, Island MD, Hausinger RP. Molecular Biology of Microbial Ureases. Microbiol Rev. 1995;59(3):451–480.

24. Konieczna I, Zarnowiec P, Kwinkowski M, et al. Bacterial Urease and its Role in Long-Lasting Human Diseases. Curr Protein Pept Sci. 2012;13(8):789–806. doi:10.2174/138920312804871094

25. Brenner DJ, O’Hara CM, Grimont PAD, et al. Biochemical identification of Citrobacter species defined by DNA hybridization and description of Citrobacter gillenii sp. nov. (formerly Citrobacter genomospecies 10) and Citrobacter murliniae sp. nov. (formerly Citrobacter genomospecies 11). J Clin Microbiol. 1999;37(8):2619–2624. doi:10.1128/jcm.37.8.2619-2624.1999

26. Davin-Regli A, Lavigne JP, Pagès JM. Enterobacter spp.: update on taxonomy, clinical aspects, and emerging antimicrobial resistance. Clin Microbiol Rev. 2019;32(4):1–32. doi:10.1128/CMR.00002-19

27. Armbruster CE, Smith SN, Johnson AO, et al. The Pathogenic Potential of Proteus mirabilis Is Enhanced by Other Uropathogens during Polymicrobial Urinary Tract Infection. Infect Immun. 2017;85(2):e00808–16.

28. Gaston JR, Andersen MJ, Johnson AO, et al. Enterococcus faecalis polymicrobial interactions facilitate biofilm formation, antibiotic recalcitrance, and persistent colonization of the catheterized urinary tract. Pathogens. 2020;9(10):1–20. doi:10.3390/pathogens9100835

29. Singh R, Rohilla RK, Sangwan K, Siwach R, Magu NK, Sangwan SS. Bladder management methods and urological complications in spinal cord injury patients. Indian J Orthop. 2011;45(2):141–147. doi:10.4103/0019-5413.77134

30. Kunin CM, Chin QF, Chambers S. Indwelling urinary catheters in the elderly. Relation of “catheter life” to formation of encrustations in patients with and without blocked catheters. Am J Med. 1987;82(3):405–411. doi:10.1016/0002-9343(87)90438-4

31. Kohler-Ockmore J, Feneley RCL. Long-term catheterization of the bladder: Prevalence and morbidity. Br J Urol. 1996;77(3):347–351. doi:10.1046/j.1464-410X.1996.09074.x

