## Supplementary Figures and Tables for "Pilot Clinical Trial to test the function of a Diagnostic Sensor in predicting Impending Urinary Catheter Blockage in Long-term Catheterized Patients"

**A.** **B.**

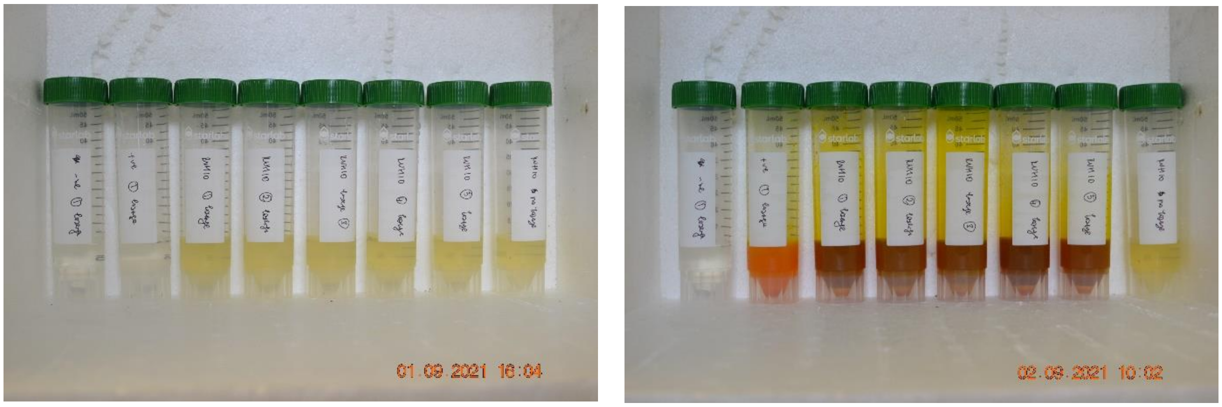


**Supplementary Figure 1.** Photograph of the tubes to test the sensors, sample RUH10. A. 0 h, B. 18 h.

**Supplementary Table 1.** Summary of the catheter manufacturers used.

| **Variable** | **Participants** |
|---|---|
| Catheter manufacture | |
| Rusch | 4 |
| Bard | 4 |
| Coloplast Coude | 4 |
| Coloplast Porges | 3 |
| Yushin | 3 |
| Brilliant AquaFlate | 2 |
| Mediplus | 2 |
| Medicath | 1 |
| Libra | 1 |
| Not recorded | 4 |

**Supplementary Table 2.** Bacterial species identified for each sample donated.

| Sample number | Species identified |
|---|---|
| **RUH01** | *Shigella flexneri* |
| | *Pseudomonas aeruginosa* |
| | *Enterococcus faecalis* |
| **RUH02** | *Shigella flexneri* |
| **RUH03** | *Pseudomonas aeruginosa* |
| **RUH04** | *Staphylococcus epidermidis* |
| **RUH05** | *Enterococcus faecium* |
| **RUH06** | *Proteus mirabilis* |
| **RUH07** | *Enterococcus faecalis* |
| | *Proteus mirabilis* |
| **RUH08** | *Brevibacterium frigoritolerans* |
| | *Proteus alimentorum* |
| **RUH09** | *Escherichia spp.* |
| | *Pseudomonas aeruginosa* |
| | *Proteus mirabilis* |
| **RUH10** | *Mammaliiococcus lentus* |
| | *Proteus mirabilis* |
| | *Staphylococcus epidermidis* |
| **RUH11** | *Escherichia spp.* |
| | *Proteus mirabilis* |
| **RUH12** | *Klebsiella michiganensis* |
| | *Proteus mirabilis* |
| **RUH13** | *Proteus mirabilis* |
| **RUH14** | *Klebisella michiganensis* |
| | *Morganella morganii* |
| **RUH15** | *Enterobacter hormaechei* |
| **RUH16** | *Enterococcus faecalis* |
| | *Proteus mirabilis* |
| **RUH17** | *Klebisella quasivariicola* |
| | *Pseudmonas aerugionosa* |
| | *Shigella flexneri* |
| **RUH18** | *Escherichia fergusonii* |
| | *Shigella flexneri* |
| **RUH19** | *Proteus mirabilis* |
| **RUH20** | *Citrobacter koseri* |
| | *Enterococcus faecalis* |
| **RUH21** | *Pseudomonas junteni* |
| | *Staphylococcus schweitzeri* |
| **RUH22** | *Klebsiella pneumoniae* |
| | *Pseudomonas aeruginosa* |

| | |
|---|---|
| **RUH23** | *Enterococcus faecalis* |
| | *Pseudomonas aeruginosa* |
| **RUH24** | *Enteroccous faecalis* |
| | *Escherichia fergusonii* |
| | *Pseudomonas aeruginosa* |
| **RUH25** | *Citrobacter koseri* |
| **RUH26** | *Proteus mirabilis* |
| **RUH27** | *Klebsiella aerogenes* |
| **RUH28** | *Enterococcus faecalis* |
| | *Enterobacter hormaechei* |
| **RUH29** | *Enterococcus faecalis* |
| | *Corynebacterium spp.* |
| **RUH30** | *Enterococcus faecalis* |
| | *Pseudomonas aeruginosa* |
| **RUH31** | *Klebsiella michiganensis* |
| | *Proteus mirabilis* |
| **RUH32** | *Klebisella michianensis* |
| | *Proteus mirabilis* |
| **RUH33** | *Staphylococcus epidermidis* |
| | *Klebisella michianensis* |
| **RUH34** | *Providencia spp.* |
| | *Klebsiella pneumoniae* |
| **RUH35** | *Pseudomonas aeruginosa* |
| | *Citrobacter koseri* |
