## Supplementary Document 1 for "Pilot Clinical Trial to test the function of a Diagnostic Sensor in predicting Impending Urinary Catheter Blockage in Long-term Catheterized Patients"

ICIQ-LTCqol 12/12

Initial number ☐☐ ☐☐ ☐☐ ☐☐☐

**CONFIDENTIAL**

DAY MONTH YEAR ☐☐ ☐☐ ☐☐
**Today's date**

### Living with a long term catheter

This form can be filled in by a carer, friend or family member, as well as, or instead of by you, but it needs to be your own opinions about your catheter (not theirs). We have used "**you**" to refer to the **person with the catheter** throughout the form – no matter who fills in the form.

**Please could you tell us who is answering the questionnaire by ticking the box:**

Catheter user ☐ 1 Carer ☐ 2 Family member/friend ☐ 3 Other ☐ 4

1. **Is the catheter user** *(tick one)***:** Female ☐ Male ☐

2. **Please write in the catheter user's date of birth:** ☐☐ ☐☐ ☐☐
DAY MONTH YEAR

**3. What type of catheter do you currently have?** *(Tick one box)*

| | | |
|---|---|---|
| urethral (between the legs/into the penis) | ☐ | 1 |
| supra-pubic (into the abdomen) | ☐ | 2 |

#### Catheter function and concern

**4a. Do you have confidence in your catheter equipment (catheter, tubing and bag)?** *(Tick one box)*

| | | |
|---|---|---|
| never | ☐ | 4 |
| occasionally | ☐ | 3 |
| sometimes | ☐ | 2 |
| most of the time | ☐ | 1 |
| all of the time | ☐ | 0 |

**4b. How much does this bother you?**
*Please ring a number between 0 (not at all) and 10 (a great deal)*

**0** 1 2 3 4 5 6 7 8 9 **10**
not at all — a great deal

**5a. Is the possibility of the catheter leaking on your mind?** *(Tick one box)*

| | | |
|---|---|---|
| never | ☐ | 0 |
| occasionally | ☐ | 1 |
| sometimes | ☐ | 2 |
| most of the time | ☐ | 3 |
| all of the time | ☐ | 4 |

**5b. How much does this bother you?**
*Please ring a number between 0 (not at all) and 10 (a great deal)*

**0** 1 2 3 4 5 6 7 8 9 **10**
not at all — a great deal

Copyright © "ICIQ Group"

**6a.** **Is the possibility of the catheter <u>blocking</u> on your mind?** *(Tick one box)*

| | | |
|---|---|---|
| never | ☐ | 0 |
| occasionally | ☐ | 1 |
| sometimes | ☐ | 2 |
| most of the time | ☐ | 3 |
| all of the time | ☐ | 4 |

**6b.** **How much does this bother you?**
*Please ring a number between 0 (not at all) and 10 (a great deal)*

**0** 1 2 3 4 5 6 7 8 9 **10**
not at all — a great deal

**7a.** **How problematic is your catheter?** *(Tick one box)*

| | | |
|---|---|---|
| problem free | ☐ | 0 |
| some problems but I would rather keep it | ☐ | 1 |
| some problems but I have to keep it | ☐ | 2 |
| lots of problems | ☐ | 3 |

**8a.** **How often do you have 'urine infections' that make you feel unwell or require you to take antibiotics?** *(Tick one box)*

| | | |
|---|---|---|
| several times per month | ☐ | 5 |
| about once a month | ☐ | 4 |
| once every 2-3 months | ☐ | 3 |
| a couple of times a year | ☐ | 2 |
| less than once a year | ☐ | 1 |
| never | ☐ | 0 |

**8b.** **How much does this bother you?**
*Please ring a number between 0 (not at all) and 10 (a great deal)*

**0** 1 2 3 4 5 6 7 8 9 **10**
not at all — a great deal

**9a.** **Does your catheter cause you to worry about smell?** *(Tick one box)*

| | | |
|---|---|---|
| never | ☐ | 0 |
| occasionally | ☐ | 1 |
| sometimes | ☐ | 2 |
| most of the time | ☐ | 3 |
| all of the time | ☐ | 4 |

**9b.** **How much does this bother you?**
*Please ring a number between 0 (not at all) and 10 (a great deal)*

**0** 1 2 3 4 5 6 7 8 9 **10**
not at all — a great deal

Copyright © "ICIQ Group"

**10a. Are you embarrassed by having a catheter?** *(Tick one box)*

| | | |
|---|---|---|
| never | ☐ | 0 |
| occasionally | ☐ | 1 |
| sometimes | ☐ | 2 |
| most of the time | ☐ | 3 |
| all of the time | ☐ | 4 |

**10b. How much does this bother you?**
*Please ring a number between 0 (not at all) and 10 (a great deal)*

**0** 1 2 3 4 5 6 7 8 9 **10**
not at all — a great deal

**11a. Do you feel you have adapted to life with a catheter?** *(Tick one box)*

| | | |
|---|---|---|
| not at all | ☐ | 4 |
| not really | ☐ | 3 |
| somewhat | ☐ | 2 |
| mostly | ☐ | 1 |
| completely | ☐ | 0 |

**11b. How much does this bother you?**
*Please ring a number between 0 (not at all) and 10 (a great deal)*

**0** 1 2 3 4 5 6 7 8 9 **10**
not at all — a great deal

**11c. Please provide further details if you would like**

**12a. Overall, how much does having a catheter affect your everyday life?**
*Please ring a number between 0 (overall having a catheter is good) and 10 (overall having a catheter is bad)*

**0** 1 2 3 4 5 6 7 8 9 **10**
overall good — overall bad

**Catheter function and concern score: sum scores 4a-12a** ☐ ☐

Copyright © "ICIQ Group"

#### Lifestyle impact

**13. Does your catheter affect your ability to travel?** *(Tick one box)*

| | | |
|---|---|---|
| I don't travel but for other reasons | ☐ | 5 |
| I don't travel because of my catheter | ☐ | 4 |
| the catheter limits my ability to travel | ☐ | 3 |
| the catheter has no effect on my ability to travel | ☐ | 2 |
| the catheter has helped my ability to travel | ☐ | 1 |

**14. Does your catheter affect your social activities (for example, going out for a meal)?** *(Tick one box)*

| | | |
|---|---|---|
| I don't take part in social activities but for other reasons | ☐ | 5 |
| I don't take part in social activities because of my catheter | ☐ | 4 |
| the catheter limits my ability to take part in social activities | ☐ | 3 |
| the catheter has no effect on my social activities | ☐ | 2 |
| the catheter has helped my ability to take part in social activities | ☐ | 1 |

**15. Does your catheter affect your ability to go out of the house?** *(Tick one box)*

| | | |
|---|---|---|
| I don't go out but for other reasons | ☐ | 5 |
| I don't go out because of my catheter | ☐ | 4 |
| the catheter limits my ability to go out | ☐ | 3 |
| the catheter has no effect on my ability to go out | ☐ | 2 |
| the catheter has helped me to go out | ☐ | 1 |

**Lifestyle impact score: sum scores 13-15** ☐ ☐

#### Unscored items

**16a. Do you use pads as well as your catheter because of your bladder?** *(Tick one box)*

| | | |
|---|---|---|
| never | ☐ | 0 |
| occasionally | ☐ | 1 |
| sometimes | ☐ | 2 |
| most of the time | ☐ | 3 |
| all of the time | ☐ | 4 |

**16b. How much does using pads because of leaks bother you?**
*Please ring a number between 0 (not at all) and 10 (a great deal)*

**0** 1 2 3 4 5 6 7 8 9 **10**

not at all — a great deal

Copyright © "ICIQ Group"

**17a.** **Does your catheter cause you any pain, discomfort or soreness?** *(Tick all that apply)*

- never ☐ 0
- occasionally ☐ 1
- sometimes ☐ 2
- most of the time ☐ 3
- always ☐ 4

**17b.** **How much does this bother you?**
*Please ring a number between 0 (not at all) and 10 (a great deal)*

**0** 1 2 3 4 5 6 7 8 9 **10**
not at all — a great deal

**18a.** **Do you experience any bladder spasm (tightening of the bladder when you don't want it to)?** *(Tick one box)*

- never ☐ 0
- occasionally ☐ 1
- sometimes ☐ 2
- most of the time ☐ 3
- always ☐ 4

**18b.** **How much does this bother you?**
*Please ring a number between 0 (not at all) and 10 (a great deal)*

**0** 1 2 3 4 5 6 7 8 9 **10**
not at all — a great deal

**19a.** **Does having a catheter prevent sexual activity?***(Tick one box)*

- never ☐ 0
- occasionally ☐ 1
- sometimes ☐ 2
- most of the time ☐ 3
- all of the time ☐ 4
- not applicable ☐ 8
- don't wish to answer ☐ 9

**19b.** **How much does this bother you?**
*Please ring a number between 0 (not at all) and 10 (a great deal)*

**0** 1 2 3 4 5 6 7 8 9 **10**
not at all — a great deal

#### Thank you very much for answering these questions.

Copyright © "ICIQ Group"
