## Supplementary Document 2 for "Pilot Clinical Trial to test the function of a Diagnostic Sensor in predicting Impending Urinary Catheter Blockage in Long-term Catheterized Patients"

**Data Protection**

In this research study we will use information from you and your medical records. We will only use information that we need for the research study. We will let very few people know your name or contact details, and only if they really need it for this study. Everyone involved in this study will keep your data safe and secure. We will also follow all privacy rules.

At the end of the study we will save some of the data in case we need to check it and for future research. We will make sure no-one can work out who you are from the reports we write. The information pack tells you more about this.

**How will we use information about you?**

We will need to use information from your medical records for this research project.
This information will include your [initials/ NHS number/ name/ contact details. People will use this information to do the research or to check your records to make sure that the research is being done properly.
People who do not need to know who you are will not be able to see your name or contact details. Your data will have a code number instead.
We will keep all information about you safe and secure.
Once we have finished the study, we will keep some of the data so we can check the results.
We will write our reports in a way that no-one can work out that you took part in the study.

**What are your choices about how your information is used?**

You can stop being part of the study at any time, without giving a reason, but we will keep information about you that we already have.
We need to manage your records in specific ways for the research to be reliable. This means that we won't be able to let you see or change the data we hold about you.
If you agree to take part in this study, you will have the option to take part in future research using your data saved from this study.

**Where can you find out more about how your information is used?**

You can find out more about how we use your information is stored and protected at

- https://www.bath.ac.uk/corporate-information/research-data-policy/
- by asking one of the research team
- by ringing us on 01225 821892

THE UROLOGY FOUNDATION

Department of Chemistry
University of Bath
Bath, BA2 7AY


The Annett Trust

Vers: 09/03/2020

### A study to test a sensor for giving early warning of catheter blockage

A team of clinicians, scientists and engineers from the Royal United Hospital in Bath and the University of Bath are developing a sensor system designed to give early warning of likely blockage of a urinary catheter, before blockage actually takes place.

The sensor is a simple 'lozenge' which would be placed in the urine drainage bag, giving a bright green colour if the pH of the urine rises above acritical level. High pH urine is indicative of bladder infection and can cause the catheter to block.

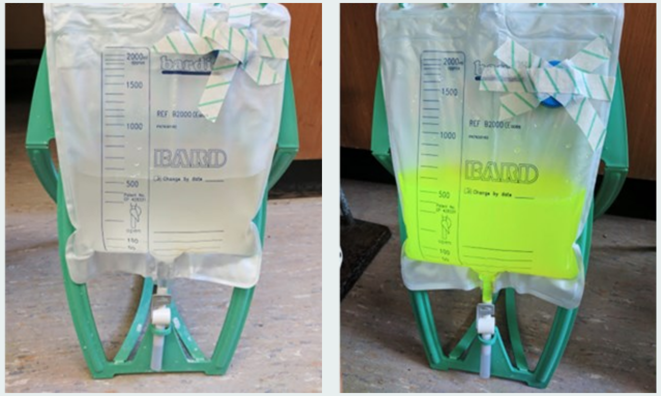

***Urine drainage bags with sensor:*** *Left is normal urine; Right the colour indicator alerts the patient of a possible urinary tract infection and to change their catheter*

This leaflet gives information about how the lozenge will look, how it will feel and how it detects infection. If you could read this leaflet and answer some questions it will really help us in developing our sensor. By taking part, you are helping us to develop an exciting new product that will be used on patients in the future. We are very grateful for your time—your opinion will directly affect how we design this sensor.

Royal United Hospitals Bath NHS
NHS Foundation Trust

UNIVERSITY OF BATH

### The Sensor

**What is it for?** Catheter blockage which is often associated with certain urinary tract bacteria can result in painful distention of the bladder, and can lead to serious illness such as acute pyelonephritis and septicaemia.

**How will it look and feel?** The sensor is a small lozenge, about 7 mm long and 3 mm wide, which will be put in the urine drainage bag (leg bag). If the pH of the urine rises, indicating possible infection, a bright green fluorescent dye will be released into the leg bag. The dye is harmless, but will not enter the bladder or get on skin. Dye release will indicate the catheter should be changed I the next 12-18 hours.

**How will it be used?** It will be put in the leg bag just prior to connecting the external drain of the catheter to the bag. Use of the sensor will not affect the feel of the catheter in any way. On colour change, the catheter should be changed within 12-18 hours.

**How does it work?** The lozenge is comprised of a special polymer which dissolves at pH greater than about 7.5 (indicative of infection). Healthy urine should be slightly acidic (pH 5.5). As the polymer dissolves, the dye inside the lozenge is released.

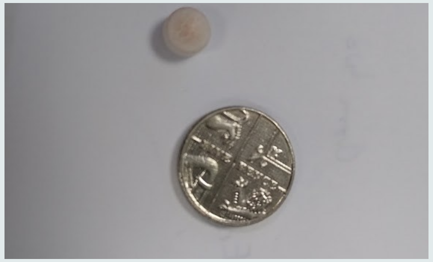

***Lozenge sensor:*** *next to a 5 pence coin (for size reference)*

### The Research Study

#### The actual study

#### The actual study

This study will not involve testing the sensor in your urine drainage bag. It will involve you consenting to donate the urine in your leg bag for the University of Bath researchers to test whether the sensor will work under laboratory conditions. The NHS Trust Urology Clinic will label the bags with your date of birth, the collection date, patient initials and a unique patient identifier number. The University of Bath researchers will not have any access to information that can identify you—you'll be completely anonymous.

We will ask you some questions about how your quality of life is affected by having a catheter using an internationally validated questionnaire, at the time of urine donation.

We will contact you approximately three weeks after donation of urine by phone to ask how you have been getting on with the catheter (whether it has been bypassing, blocking or you have needed antibiotics).

If this pilot study is successful, the clinical team might ask whether you would be interested in testing the sensor in a follow up project, probably in late 2020. You are under no obligation to agree to this, and you would be able to change your mind at any time.

#### We have some questions about this study for you:

- Do you understand why we need to do this study and what it involves?
- Do you have any questions about the study?
- If you were invited to take part, would you participate?
- If you would choose not to participate, what would your reservations be?
