## Supplementary Document 3 for "Pilot Clinical Trial to test the function of a Diagnostic Sensor in predicting Impending Urinary Catheter Blockage in Long-term Catheterized Patients"

| | | | | |
|---|---|---|---|---|
| Royal United Hospitals Bath NHS Foundation Trust<br>**Urinostics - Pilot study**<br>University of Bath | **Study ID** | R | U | H |
| | **Assessment Date** | DD | MM | YYYY |

### Registration

| | | | |
|---|---|---|---|
| 1. | Date of birth | __/__/____ (dd/mm/yyyy) | |
| 2. | Is there a suspected infection? | ☐ | Yes |
| | | ☐ | No |
| 3. | Is patient considered eligible to take part in the study? | ☐ | Yes |
| | | ☐ | No |
| 4. | Has patient signed the consent form? | ☐ | Yes |
| | | ☐ | No |

| Study ID | R | U | H |
|---|---|---|---|
| Assessment Date | DD | MM | YYYY |

### 1. Eligibility Checklist

| Inclusion Criteria | | Yes | No |
|---|---|---|---|
| 5. | Patient attending Urology Clinic | ☐ | ☐ |
| 6. | Adult aged >18 years | ☐ | ☐ |
| 7. | Patients with long term in-dwelling urinary catheters | ☐ | ☐ |
| 8. | Consent gained for study | ☐ | ☐ |
| Exclusion Criteria | | | |
| 9. | Adult without mental capacity to consent | ☐ | ☐ |
| 10. | Consent not gained for study | ☐ | ☐ |

### Patient

| | | | | | |
|---|---|---|---|---|---|
| 11. | Version of consent form | ☐.☐ | | | |
| 12. | Date signed | __/__/____ (dd/mm/yyyy) | | | |
| 13. | The patient has initialled all the boxes on the consent form | ☐ | Yes | ☐ | No |
| 14. | The patient has personally signed and dated the consent form | ☐ | Yes | ☐ | No |
| 15. | The person taking consent has signed and dated the form on the same date as the patient | ☐ | Yes | ☐ | No |
| 16. | The person taking consent has been delegated this role on the delegation log | ☐ | Yes | ☐ | No |

### Co-Morbidities

| | | | | | |
|---|---|---|---|---|---|
| 17. | Any significant co-morbidities? | ☐ | Yes | ☐ | No |
| **18.** | Co-morbidities - specify (e.g. advanced bladder cancer/medication that may alter the urine colour) Provide details: | *Provide details* | | | |
| **19.** | Has the patient experienced a catheter blockage event within the last 4 weeks? | ☐ | Yes | ☐ | No |
| **20.** | If the patient did experience a catheter blockage, please give approximate data | Date of blockage: | | | |

### Demographics

| | | | |
| --- | --- | --- | --- |
| 21. | Participant sex | ☐ | Male |
| | | ☐ | Female |
| 22. | Ethnicity | ☐ | White: British |
| | | ☐ | White: Irish |
| | | ☐ | White: Other |
| | | ☐ | Mixed/Multiple ethnic groups: White & Black Caribbean |
| | | ☐ | Mixed/Multiple ethnic groups: White & Black African |
| | | ☐ | Mixed/Multiple ethnic groups: White & Asian |
| | | ☐ | Mixed/Multiple ethnic groups: Other |
| | | ☐ | Indian |
| | | ☐ | African |
| | | ☐ | Bangladeshi |
| | | ☐ | Pakistani |
| | | ☐ | Chinese |
| | | ☐ | Other Asian background |
| | | ☐ | Black/African/Caribbean/Black British: Caribbean |
| | | ☐ | Black/African/Caribbean/Black British: African |
| | | ☐ | Black/African/Caribbean/Black British: Other |
| | | ☐ | Arab |
| | | ☐ | Other ethnic group |
| | | ☐ | *Unknown* |
| 23. | Is patient a smoker? | ☐ | Yes |
| | | ☐ | No |
| | | ☐ | *Unknown* |

| | | | |
|---|---|---|---|
| Study ID | R | U | H |
| Assessment Date | DD | MM | YYYY |

### Medications

| | | | |
|---|---|---|---|
| 24. | Any current/recent antibiotic use, within the last two weeks? | ☐ | Yes |
| 25. | Which antibiotics are being taken? | *Provide details:* | |
| 26. | Any other medication | ☐ | Riboflavin (Vit B2) |
| | | ☐ | Rifampicin |
| | | ☐ | Phenazopyridine |
| | | ☐ | Deferoxamine |
| | | ☐ | Hydroxocobalamin (Vit B12) |
| | | ☐ | Propofol |
| | | ☐ | Doxycycline |
| | | ☐ | Acetaminophen overdose |
| | | ☐ | |
| | | ☐ | Metronidazole |
| | | ☐ | Nitrofurantoin |
| | | ☐ | Other - *Provide details:* |

### Recent diet

| | | |
|---|---|---|
| **27.** | Eaten in the last 24 hrs any beetroot/asparagus/blackberries? | *Provide details:* |

### Previous catheter change

| | | | |
|---|---|---|---|
| 28. | Date | __/__/____ (dd/mm/yyyy) | |
| 29. | Reason | ☐ | Routine |
| | | ☐ | Emergency |
| **30.** | Any Farcofill added? | ☐ | Yes |
| | | ☐ | No |

| | | |
|---|---|---|
| 31. | Have any bladder washouts (with saline) been carried out between now and the last catheter change? | ☐ Yes |
| | | ☐ No |
| 32. | Have any bladder maintenance solutions (such as citric acid based solution) been used between now and the last catheter change? | ☐ Yes |
| | | ☐ No |

### Samples

| | | | |
|---|---|---|---|
| 33. | Type of Drainage bag | ☐ | Leg bag |
| | | ☐ | Drainage bag on stand |
| | | Brand | |
| | | ☐ | Other |
| 34. | Appearance of Urine in Drainage Bag | ☐ | Clear |
| | | ☐ | Cloudy |
| | | ☐ | Offensive |
| | | ☐ | Other i.e. colour (specify) |
| 35. | Type of catheter | Silicon | Yes ☐ |
| | | | No ☐ |
| | | Size | |
| | | Brand | |
| 36. | Appearance of Catheter | ☐ | Clean |
| | | ☐ | Cloudy |
| | | ☐ | Encrusted |
| | | ☐ | Other (specify) |
| 37. | Were the urinary drainage bag and catheter placed in separate labelled storage bags? | ☐ | Yes |
| | | ☐ | No |
| 38. | Date of urine collection | __/__/____ (dd/mm/yyyy) | |
| 39. | Time of urine collection (24 hour clock) | ___:___ | |
| 40. | Were the bags incubated at 2 - 8 °C in the clinical area where the patient is being treated? | ☐ Yes | ☐ No |

| | | | | | |
|---|---|---|---|---|---|
| 41. | Has the patient been previously recruited to this study? | ☐ | Yes | ☐ | No |
| 42. | If yes to Q41, please give previous ID number: (refer to Urinostics site subject list, Patient information, divider 3 in site file) | **Study ID** | | | |
