## Supplementary Document 4 for "Pilot Clinical Trial to test the function of a Diagnostic Sensor in predicting Impending Urinary Catheter Blockage in Long-term Catheterized Patients"

| | | | | | | |
|---|---|---|---|---|---|---|
| Urinostics study | Study ID | R | U | H | | |
| | Assessment Date | DD | | MM | | YYYY |

### Follow-up telephone questionnaire

| | | | |
|---|---|---|---|
| 1. | Date of birth | __/__/____ (dd/mm/yyyy) | |
| 2. | What was the date of your last catheter change? | __/__/____ (dd/mm/yyyy) | |
| 3. | Was this catheter change a routine change? | ☐ | Yes |
| | | ☐ | No |
| 4. | If the catheter change was an emergency, why was the catheter changed? | | |
| 5. | Since your last catheter change have you used any bladder washouts or maintenance solutions? | ☐ | Yes |
| | | ☐ | No |
| 6. | If you have used maintenance solutions, which ones have you used? | | |
| 7. | Have you been prescribed any antibiotics to treat a catheter-associated urinary tract infection? If so, please say which ones. | ☐ | Yes |
| | | ☐ | No |
| | | Antibiotics prescribed: | |
| 8. | Is their any feedback you want to give regarding the clinical trial? | | |

Version: 1.0 10/10/2019
