## Supplementary figures and images for "Pilot Clinical Trial to test the function of a Diagnostic Sensor in predicting Impending Urinary Catheter Blockage in Long-term Catheterized Patients"

### Supplementary Document 5

$10^3$

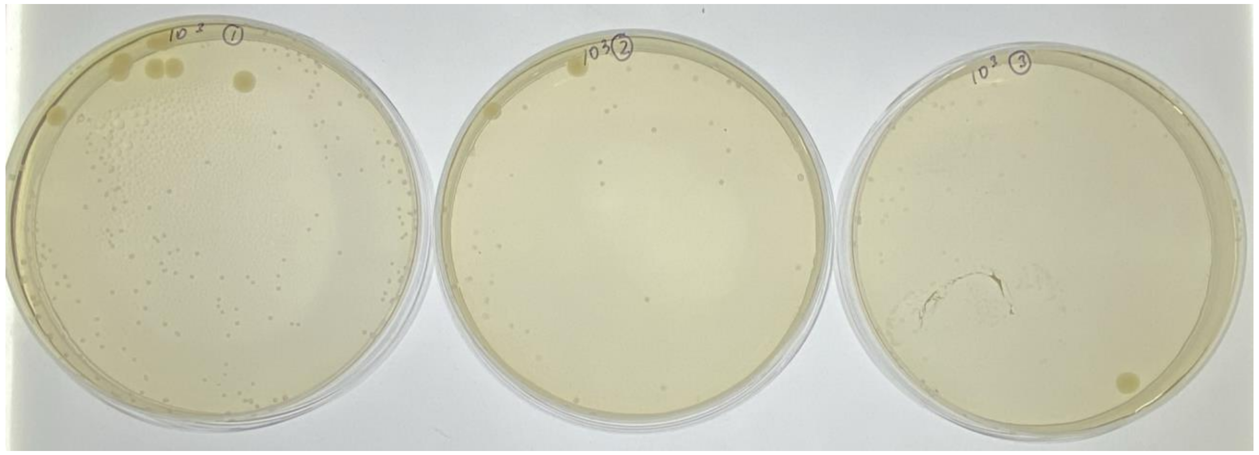

$10^4$

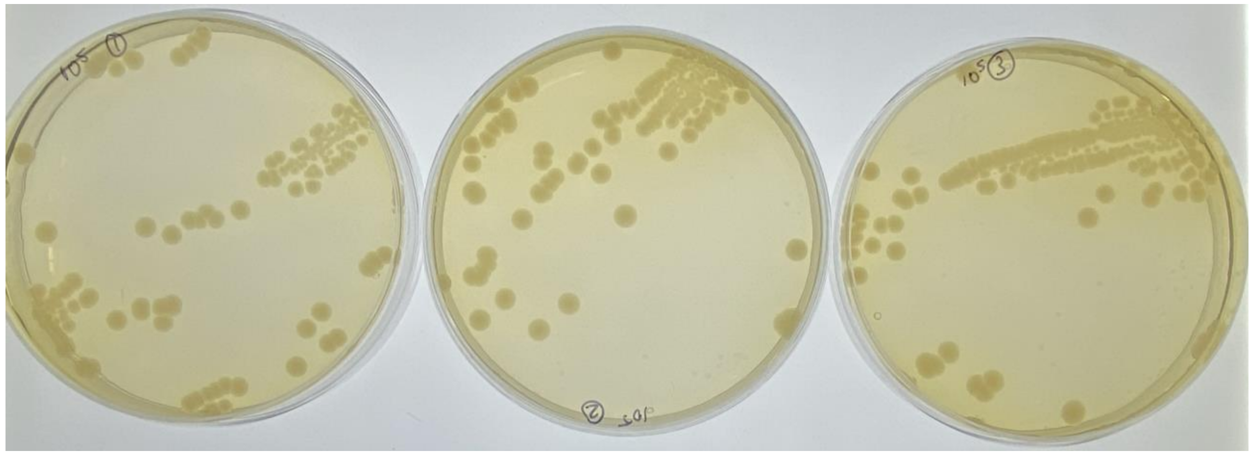

$10^5$

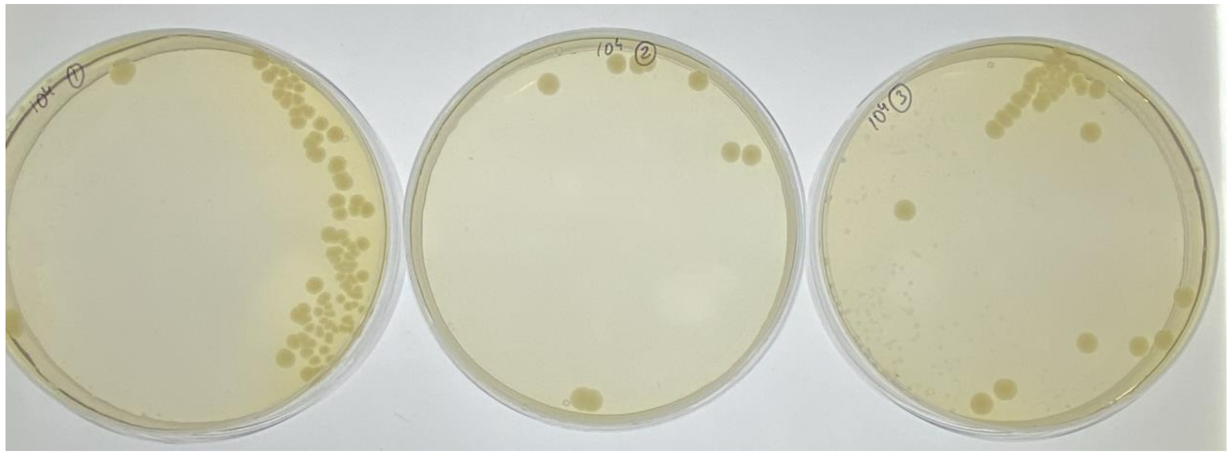

$10^6$

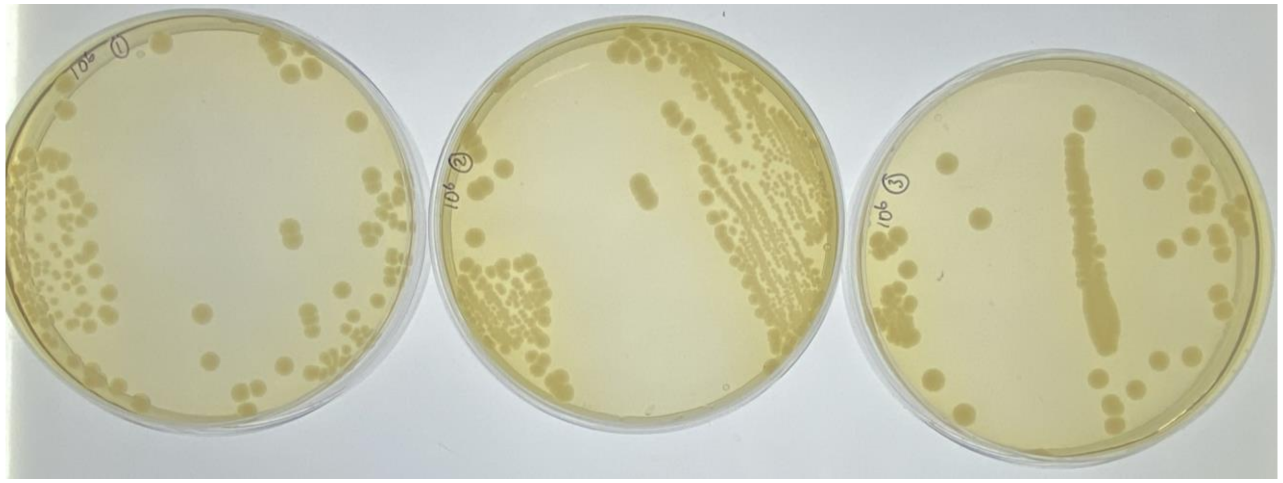

$10^7$

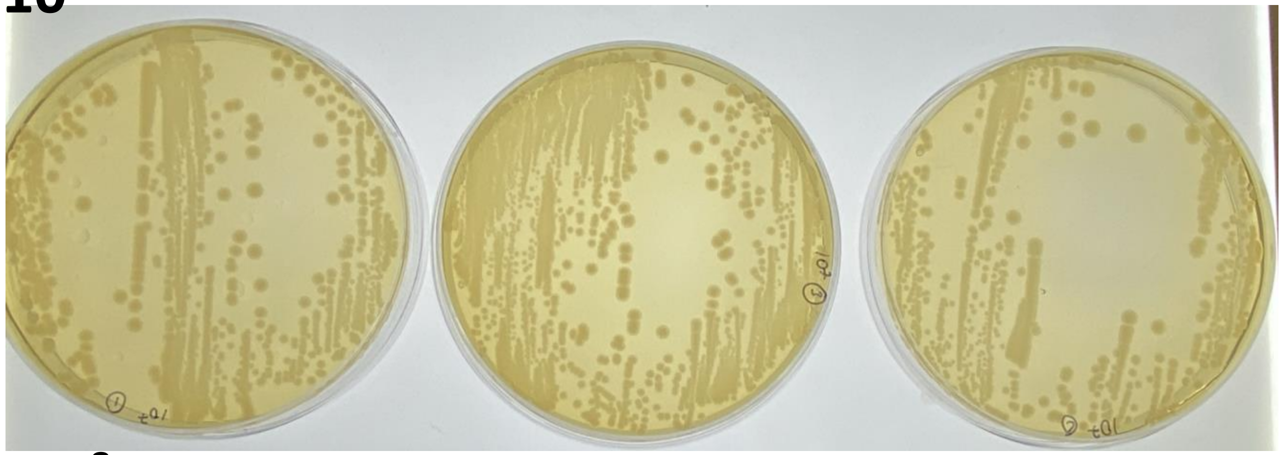

$10^8$

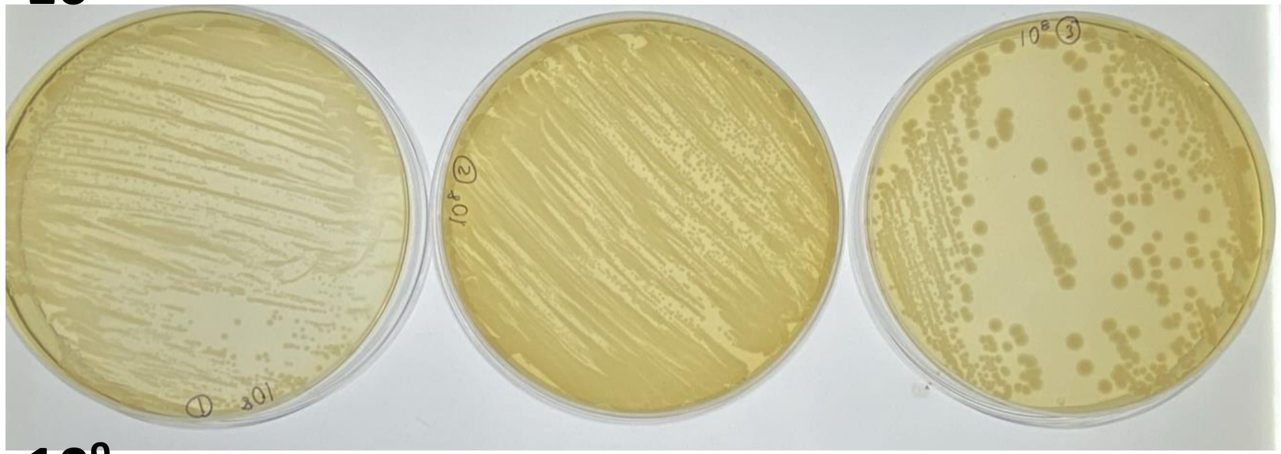

$10^9$

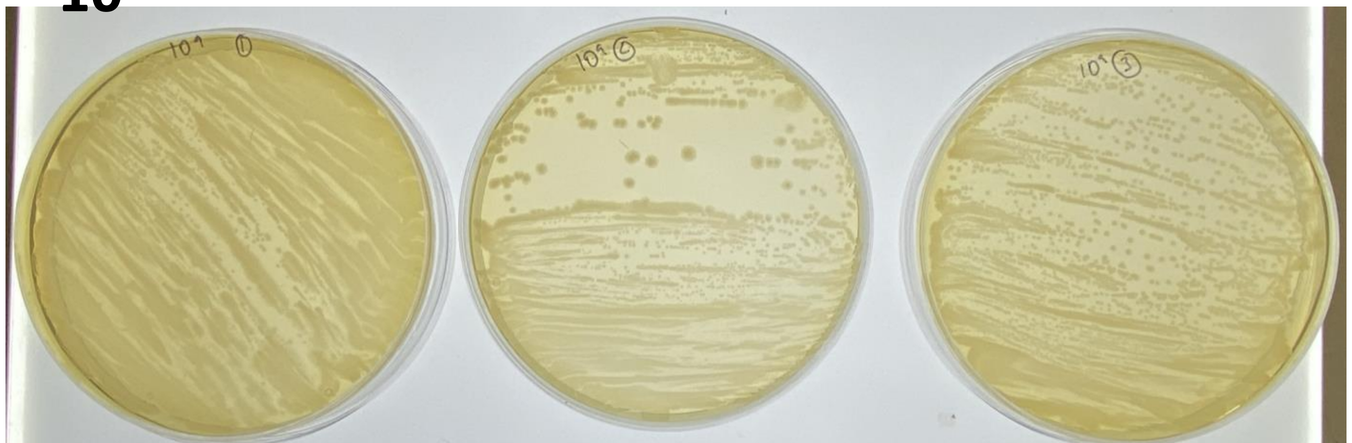
